# SARS-CoV-2 virus dynamics in recently infected people – data from a household transmission study

**DOI:** 10.1101/2022.03.17.22272516

**Authors:** Alexandra M. Mellis, Jennifer K Meece, Natasha B. Halasa, James D. Chappell, Huong Q. McLean, Carlos G. Grijalva, Kayla E. Hanson, Yuwei Zhu, Ahra Kim, Jessica Deyoe, Lynn C. Ivacic, Carrie Reed, H. Keipp Talbot, Melissa A. Rolfes

## Abstract

We used daily real-time reverse-transcription polymerase chain reaction (rRT-PCR) results from 67 cases of SARS-CoV-2 infection in a household transmission study to examine the trajectory of cycle threshold (Ct) values, an inverse correlate of viral RNA concentration, from nasal specimens collected between April 2020 and May 2021. Ct values varied over the course of infection, across RT-PCR platforms, and by participant age. Specimens collected from children and adolescents showed higher Ct values and adults aged ≥50 years showed lower Ct values than adults aged 18-49 years. Ct values were lower on days when participants reported experiencing symptoms.

## Introduction

Cycle threshold (Ct) values are generated from real-time reverse-transcription polymerase chain reaction (RT-PCR) assays and represent the minimum number of amplification cycles needed to generate a signal for a specific target. Ct values are sometimes used as a surrogate signal for SARS-CoV-2 viral loads[1], as they are inversely related to the amount of virus present in the tested specimen, but are best interpreted as viral RNA template concentration. Widespread availability has led to comparisons of Ct values at the patient[2] and community[3] levels to infer associations with illness severity and patient characteristics.

However, Ct values can vary by assay, specimen type and quality, and time during the infection course, especially complicating cross-sectional comparisons. Use of serial specimens collected from one individual over the course of infection on the same assay can partially mitigate these concerns, yet few investigations have used serial sampling to describe the natural history of SARS-Co-V2 infection[4, 5] and even fewer have been conducted among the general population[6, 7].

We described SARS-CoV-2 RT-PCR Ct values in newly infected individuals who underwent daily specimen collection as part of a multi-site prospective household transmission study. We examined the impact of age and symptoms on Ct value trajectories to understand the natural history of infection.

## Methods

We conducted a household transmission study of SARS-CoV-2 in Tennessee and Wisconsin[8] between April 2020 and May 2021. Index participants were first identified as non-hospitalized individuals who had tested positive for SARS-CoV-2 by a provider-ordered nucleic acid amplification test, and resided with at least one other individual. Index participants and household contacts were enrolled and consented within 6 days of symptom onset in the index participant. Study procedures included daily swabbing and symptom diaries (assessing presence or absence of 16 symptoms, see Supplementary Table 1), for 14 consecutive days. All participants also completed demographic surveys and self-reported pre-existing conditions (asthma, chronic liver disease, premature birth, cardiac conditions, diabetes, cancer, immunocompromising conditions, extreme obesity, kidney disease, or pregnancy) and, after COVID-19 vaccines became available, vaccination status (which was also verified against health records and immunization information systems).

Anterior nasal swabs were self-/parent-collected by study participants. Swabs were either placed in viral transport media (Remel MicroTest M4RT ®, Lenexa, KS USA) and refrigerated by participants for 7-10 days before transport, or placed in inactivating viral transport media (Primestore®, Longhorn Vaccines & Diagnostics LLC, Bethesda, MD) and stored at room temperature by participants for 1-3 days before transport to Marshfield Clinic Research Institute or Vanderbilt University Medical Center laboratories for processing and freezing at −80°C prior to testing. All specimens were tested by RT-PCR using either the CDC 2019-Novel Coronavirus Real-Time RT-PCR Diagnostic Panel (EUA CDC-006-00019; with N1 and N2 gene targets and RNase P control; CDC assay) or the ThermoFisher TaqPath™ COVID-19 ComboKit (with S and N gene and ORF1ab targets and MS2 spike control; ThermoFisher assay). Only Ct values from tests interpreted as positive (at least two SARS-CoV-2 target Ct values under 40) were analyzed. As an additional quality control measure for this analysis, we excluded Ct values from any test where the control (RNase P or MS2) result was interpreted as negative or where the RNase P control target had a Ct value greater than 35 (though this exclusion did not change results).

Viral culture was conducted on positive specimens from a subset of participants tested using the CDC assay. Wells were seeded with Vero E6-TMPRSS2 cells, to which 100μl of participant specimen was added. Wells were monitored daily for culture positivity for five days after inoculation. If >20% of cells were detached in wells exhibiting viral cytopathic effect, the specimen was interpreted as culture positive. Additional detail of culture methods and results are included in another report[9].

To examine trajectories of Ct values over the course of infection, we selected data from household contacts who met the following criteria: individuals ‘ first specimen must have been negative, they must have tested positive in 3 or more tests, and all tests must have been conducted on the same assay (Supplemental Figure 1). Days of lowest Ct values were defined per target (N1, N2, N, S, or ORF1ab). We described Ct values over time using generalized additive models controlling for the target of each assay (which also differed by assay type), with a random effect spline for repeated measurements. We compared three models for age: 1) a null model with no age effect and a single smoothing spline for the effect of each day since first testing positive, representing a null hypothesis; 2) a model with categorical age (0-11, 12-17, 18-49, or ≥50 years) and a single spline for time since first positive specimen, representing an impact of age on Ct values but not necessarily on how those Ct values change over time; 3) a model with both age and different splines for time since first positive in each age category, representing an impact of age both on Ct values overall and on how trajectories change over time. Model 2 had the lowest Akaike Information Criterion (AIC), and accounted for the greatest deviance (50.4%). We also explored the effects of symptoms on each day of infection, controlling for age as in Model 2; symptoms were considered as binary (whether the participant reported any symptom on that day, yes/no) and categorical indicators (whether a participant was asymptomatic [never reported the symptom], pre-symptomatic, within symptomatic period, or post-symptomatic that day); the binary model was chosen by smaller AIC and used for both the primary results (on impact of any symptom) and post-hoc analysis of individual symptoms (Supplemental Table 1). Similar models were used for analysis by other demographic factors.

## Results

A total of 577 household contacts from 302 households were enrolled in the parent study April 2020 - May 2021. Sixty-seven contacts from 50 households met our criteria for “incident cases “ (52.2% male; 82.1% non-Hispanic White; 19.4% with at least one underlying condition; 92.5% symptomatic; 26.8% aged 0-11, 16.4% aged 12-17, 40.3% aged 18-49, 16.4% aged ≥50; 10.4% having received only one dose of an mRNA COVID-19 vaccine before enrollment; Table 1 for demographics and Supplemental Figure 1 for inclusion/exclusion). Associations between other demographics and Ct values are presented in Supplemental Table 3. Participants tested positive for SARS-CoV-2 for a median of 10 days (interquartile range [IQR]: 8, 12 days, although 58% of participants ‘ last specimen collected were still positive for SARS-CoV-2 and participants were tested for a median of 10 days following first positivity) and were symptomatic for a median of 10 (IQR: 7, 13) days during follow-up. The median number of days from symptom onset among incident cases to their first positive test was 0 (IQR: −1, 3) days, with symptom onset preceding first positivity in 48% of symptomatic cases (see Supplemental Figure 2 for the proportion of participants who experienced symptoms prior to positivity for each individual symptom). The median time from symptom onset to lowest Ct value was 4 (IQR: 2, 6) days, indicating that symptom onset preceded lowest Ct value. The median time from first testing positive to lowest Ct values among incident cases was 3 days (IQR: 2, 4 days). Among symptomatic cases, the median time from symptom onset to lowest Ct value was 4 (IQR: 2, 6) days. Among the 93 specimens (from 13 participants, all culture positive at least once) that underwent attempted culture, Ct values were lower in culture-positive specimens (median N1 Ct value, 26.9 [IQR: 25.0, 30.0]; median N2 Ct value, 28.3 [IQR: 26.0, 30.7] from 63 specimens) than in culture-negative specimens (median N1 Ct value, 35.6 [IQR: 34.1, 38.5]; median N2 Ct value, 38.0 [IQR: 34.9, 39.0] from 30 specimens; Wilcox test p < .001).

**Table 1.**
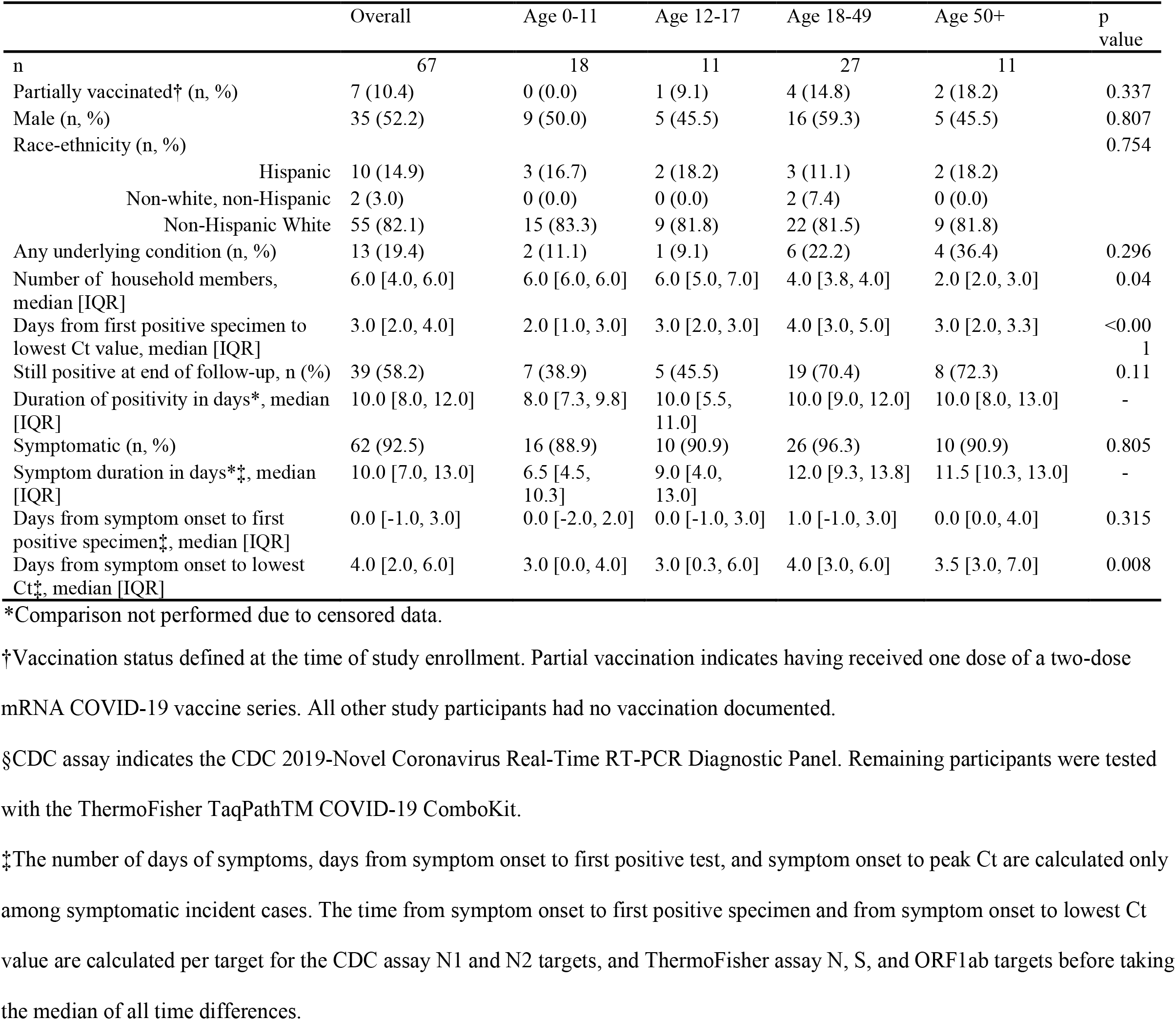
Characteristics of 67 incident cases of SARS-CoV-2 infection

|  | Overall | Age 0-11 | Age 12-17 | Age 18-49 | Age 50+ | p value |
| --- | --- | --- | --- | --- | --- | --- |
| n | 67 | 18 | 11 | 27 | 11 |  |
| Partially vaccinated† (n, %) | 7 (10.4) | 0 (0.0) | 1 (9.1) | 4 (14.8) | 2 (18.2) | 0.337 |
| Male (n, %) | 35 (52.2) | 9 (50.0) | 5 (45.5) | 16 (59.3) | 5 (45.5) | 0.807 |
| Race-ethnicity (n, %) |  |  |  |  |  | 0.754 |
| Hispanic | 10 (14.9) | 3 (16.7) | 2 (18.2) | 3 (11.1) | 2 (18.2) |  |
| Non-white, non-Hispanic | 2 (3.0) | 0 (0.0) | 0 (0.0) | 2 (7.4) | 0 (0.0) |  |
| Non-Hispanic White | 55 (82.1) | 15 (83.3) | 9 (81.8) | 22 (81.5) | 9 (81.8) |  |
| Any underlying condition (n, %) | 13 (19.4) | 2 (11.1) | 1 (9.1) | 6 (22.2) | 4 (36.4) | 0.296 |
| Number of household members, median [IQR] | 6.0 [4.0, 6.0] | 6.0 [6.0, 6.0] | 6.0 [5.0, 7.0] | 4.0 [3.8, 4.0] | 2.0 [2.0, 3.0] | 0.04 |
| Days from first positive specimen to lowest Ct value, median [IQR] | 3.0 [2.0, 4.0] | 2.0 [1.0, 3.0] | 3.0 [2.0, 3.0] | 4.0 [3.0, 5.0] | 3.0 [2.0, 3.3] | <0.001 |
| Still positive at end of follow-up, n (%) | 39 (58.2) | 7 (38.9) | 5 (45.5) | 19 (70.4) | 8 (72.3) | 0.11 |
| Duration of positivity in days*, median [IQR] | 10.0 [8.0, 12.0] | 8.0 [7.3, 9.8] | 10.0 [5.5, 11.0] | 10.0 [9.0, 12.0] | 10.0 [8.0, 13.0] | - |
| Symptomatic (n, %) | 62 (92.5) | 16 (88.9) | 10 (90.9) | 26 (96.3) | 10 (90.9) | 0.805 |
| Symptom duration in days*‡, median [IQR] | 10.0 [7.0, 13.0] | 6.5 [4.5, 10.3] | 9.0 [4.0, 13.0] | 12.0 [9.3, 13.8] | 11.5 [10.3, 13.0] | - |
| Days from symptom onset to first positive specimen‡, median [IQR] | 0.0 [-1.0, 3.0] | 0.0 [-2.0, 2.0] | 0.0 [-1.0, 3.0] | 1.0 [-1.0, 3.0] | 0.0 [0.0, 4.0] | 0.315 |
| Days from symptom onset to lowest Ct‡, median [IQR] | 4.0 [2.0, 6.0] | 3.0 [0.0, 4.0] | 3.0 [0.3, 6.0] | 4.0 [3.0, 6.0] | 3.5 [3.0, 7.0] | 0.008 |

\*Comparison not performed due to censored data.
†Vaccination status defined at the time of study enrollment. Partial vaccination indicates having received one dose of a two-dose mRNA COVID-19 vaccine series. All other study participants had no vaccination documented.
§CDC assay indicates the CDC 2019-Novel Coronavirus Real-Time RT-PCR Diagnostic Panel. Remaining participants were tested with the ThermoFisher TaqPath™ COVID-19 ComboKit.
‡The number of days of symptoms, days from symptom onset to first positive test, and symptom onset to peak Ct are calculated only among symptomatic incident cases. The time from symptom onset to first positive specimen and from symptom onset to lowest Ct value are calculated per target for the CDC assay N1 and N2 targets, and ThermoFisher assay N, S, and ORF1ab targets before taking the median of all time differences.

A total of 544 specimens were analyzed, including 1384 Ct values against SARS-CoV-2 targets. Supplemental Table 2 reports Ct values by target and age, with sample sizes of participants and tests. On average, children aged 0-11 years had Ct values that were 3.5 units higher than adults aged 18-49 years (95% confidence interval [CI]: 2.8, 4.1; p < 0.001).

Adolescents aged 12-17 years also had higher average Ct values (absolute difference: 2.7; [CI: 1.9, 3.4]; p < 0.001) and older adults, aged ≥50 years, had significantly lower Ct values (absolute difference: −1.7; [CI: −2.4, −1.0]; p < 0.001) compared with adults aged 18-49 years (Figure 1). As expected, Ct values differed between assays (higher in the CDC assay).

**Figure 1.**
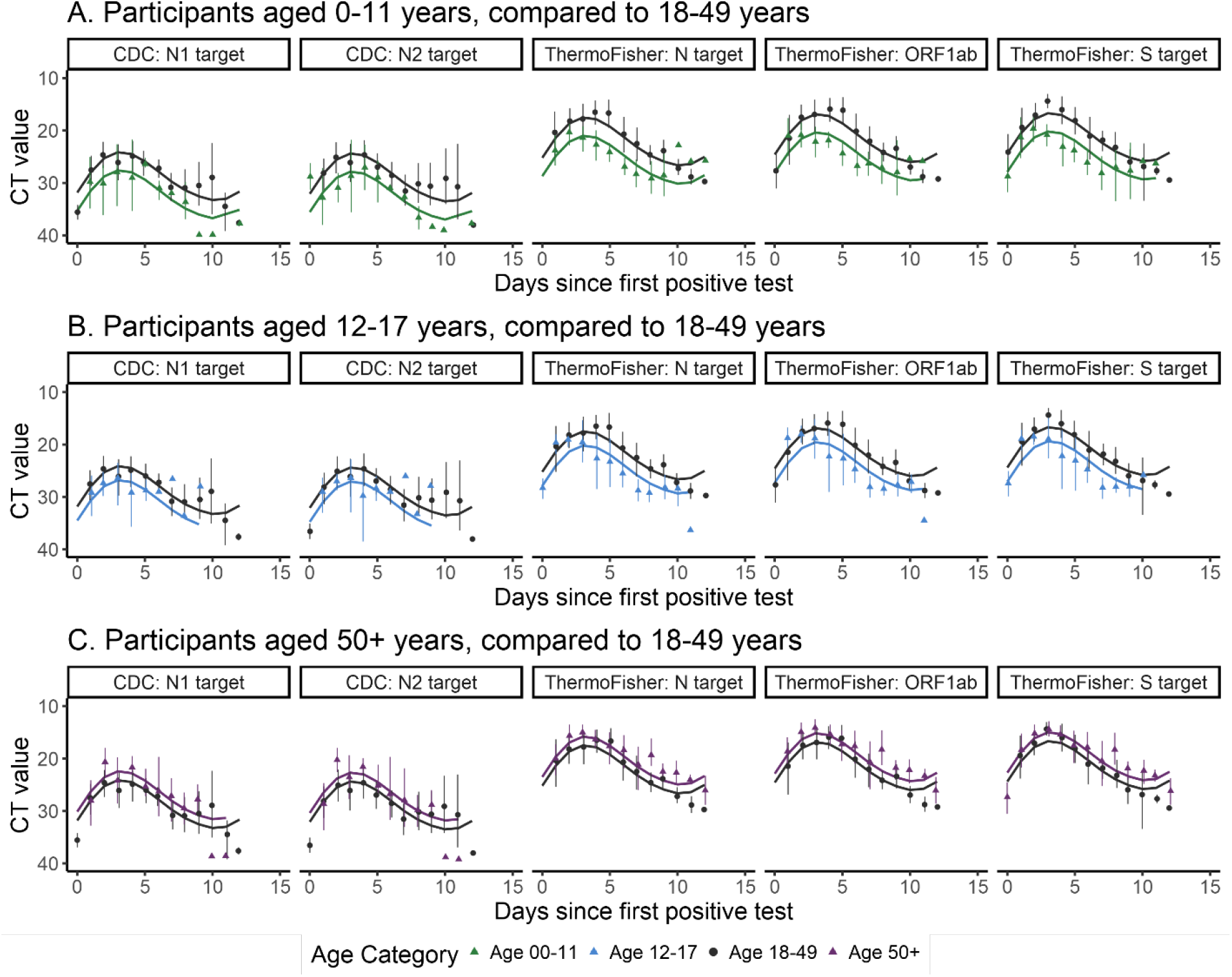
Ct value curves over time since each participant first tested positive against each target, within age groups. Dots represent mean observed values within age groups, and vertical bars show bootstrapped 95% confidence intervals. Smooth lines represent predicted values from the Generalized Additive Model of Ct values over time, accounting for age and repeated measurements. Panel A shows results from participants age 0-11 (triangle) compared to the reference group, age 18-49 (circle); Panels B and C repeat this comparison with age 12-17 or 50+. Each plot from left to right represents a SARS-CoV-2 target from one of the two included testing platforms (CDC 2019-Novel Coronavirus Real-Time RT-PCR Diagnostic Panel or the ThermoFisher TaqPathTM COVID-19 ComboKit).

Reporting symptoms on a given day was associated with lower daily Ct values, controlling for both target and age (absolute difference: −0.84 [CI: −1.0, −0.7], p < 0.001). Post-hoc tests (Supplemental Table 1) showed that Ct values were significantly lower on days participants reported having fatigue, fever, aches, chills, diarrhea, cough, chest tightness/pain, shortness of breath, wheezing, nasal congestion, runny nose, sore throat, or headache. No significant difference in Ct values were noted on days the participants experienced abdominal pain, vomiting, or loss/change of taste/smell.

## Discussion

Using data from incident cases from an intensive, prospective household study, this report contributes data on Ct values early in the infection period, which are difficult to capture using other designs. Compared to adults aged 18-49 years, we observed that Ct values were higher among children and adolescents (0-11 and 12-17 years; reflective of lower RNA levels), and lower among older adults (≥50 years) in this largely pre-Alpha-predominant period. These results are consistent with previous findings of variable Ct values by RT-PCR assay and time course of infection [10]. Other studies have reported differences in Ct values across individuals who were persistently asymptomatic[7, 11], and this analysis contributes that daily symptom status (and not just overall symptom presentation) is associated with daily Ct values, with lower Ct values on days when participants experienced symptoms. Furthermore, dates of symptom onset preceded dates of lowest Ct values by a median of 4 days, suggesting relatively longer periods of rising viral RNA concentration following symptom onset compared to other studies[4, 7].

While other studies have reported significant differences in Ct values as a function of age, the direction and interpretation of these results has differed. Cross-sectional and retrospective studies examining Ct values among children have observed lower Ct values in children under the age of 5 compared to adults over age 18[12] and compared to older children between age 5 and 14[13]. In this analysis of specimens collected daily since the first positive test result, Ct values among children and adolescents were higher than values among adults aged 18-49 years. The discrepancies between these findings and prior reports merit further investigation, and may have been driven by differences in the severity of illness, the time during infection, circulating variants, or the prospective versus cross-sectional study design.

Describing dynamics of Ct values based on frequent, systematic sampling of individuals over time ameliorates multiple concerns with the use of this data; however, these findings must still be interpreted with caution. Our selection of incident cases may have biased the sample towards those exhibiting delayed replication. Sample size and study period are also limitations, especially in our ability to assess the impact of vaccination status or dissociate vaccination or other demographics from age. Ct values also cannot be converted to a quantitative representation of viral load, and cannot be used to directly infer differences in infectiousness. However, despite these limitations, clinical interpretation of Ct values (or their trajectories) may be “tempting “ (*IDSA and AMP joint statement on the use of SARS-CoV-2 PCR cycle threshold (Ct) values for clinical decision-making*, page 3) [14]. The present data are directly relevant to these interpretations. Specifically, specimens that were collected within 4 days of symptom onset may represent periods when Ct values are still declining.

These findings contribute to our understanding of RT-PCR Ct values during SARS-CoV-2 infections over a broad range of ages, in a community setting with mostly mildly symptomatic illness, and among individuals with a known date of first shedding. While these data were collected prior to Delta and Omicron circulation, and prior to widespread vaccination, they may provide context for interpreting trajectories in Ct values in similar populations during later SARS-CoV-2 outbreaks.

## Data Availability

Data produced in the present study may be available upon reasonable request to the authors.

## Author Note

The findings and conclusions in this report are those of the authors and do not necessarily represent the official position of the US Centers for Disease Control and Prevention.

## Ethics statement

The study protocol was approved by Institutional Review Boards at Vanderbilt University Medical Center and Marshfield Clinic Research Institute. CDC determined this activity was conducted consistent with applicable federal law and CDC policy (see 45 C.F.R. part 46; 21 C.F.R. part 56).

