## Supplementary material for "SARS-CoV-2 virus dynamics in recently infected people – data from a household transmission study"

### Supplement

Figure 1. Flow diagram

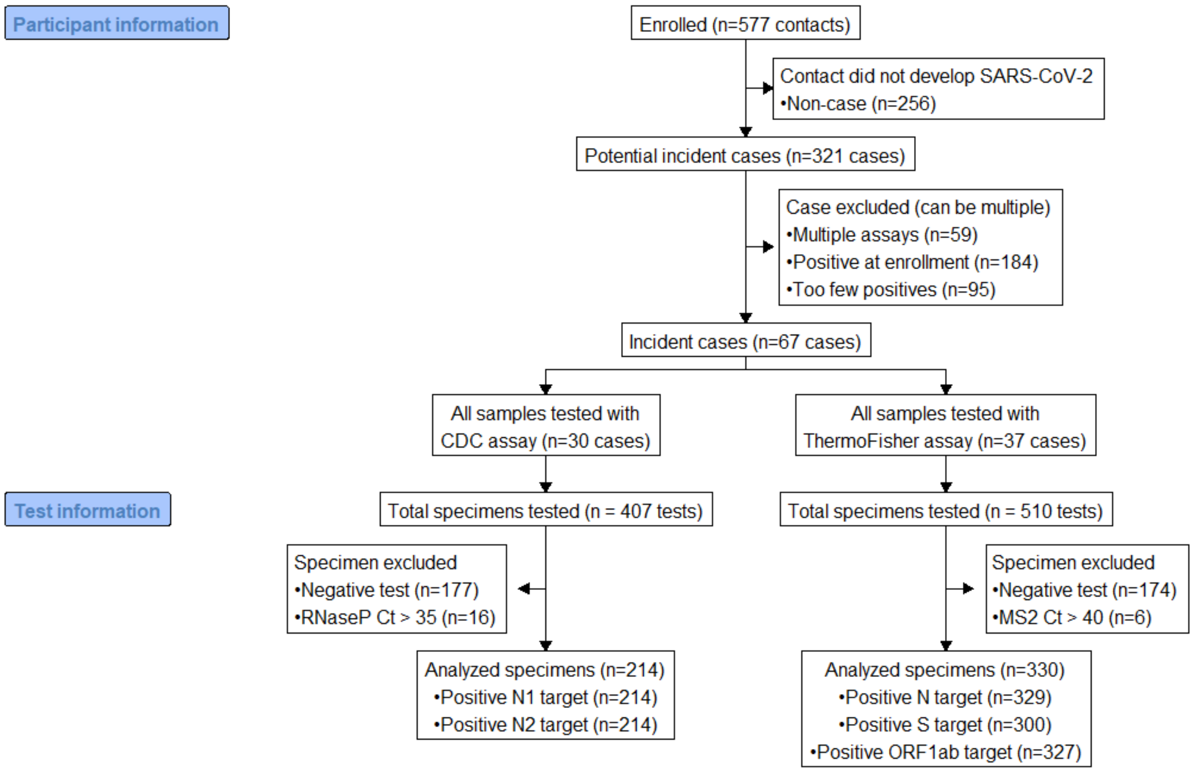


Supplemental Figure 1 shows the flow diagram for inclusion of participants and tests in this analysis. Here, CDC assay indicates CDC 2019-Novel Coronavirus Real-Time RT-PCR Diagnostic Panel and ThermoFisher assay indicates ThermoFisher TaqPath™ COVID-19 ComboKit. Participants were tested on one of these platforms based on testing availability at the enrolling site at the time of that participants' enrollment, and were only included in the analysis if all specimens they collected were tested using the same assay.

Figure 2. Order of dates of symptom onset and dates of first positive specimen among participants experiencing each symptom.

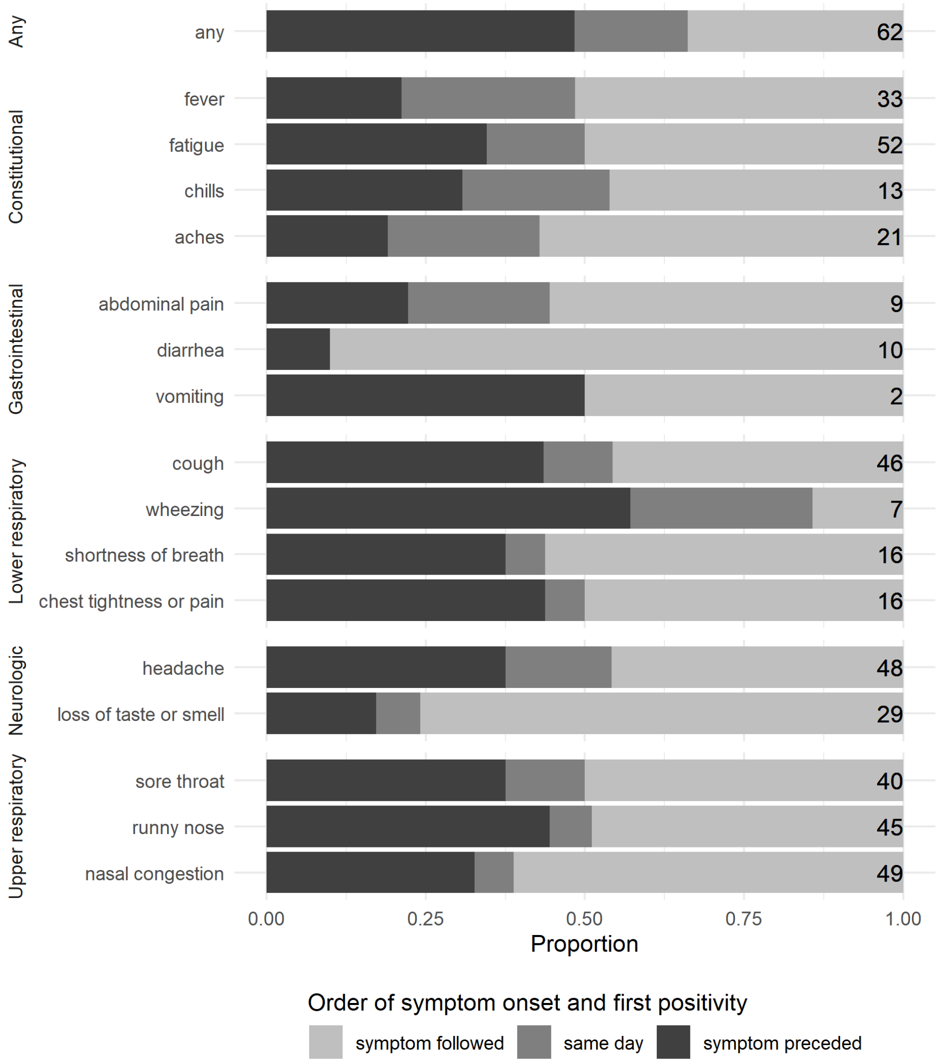


Supplemental Figure 2 shows the order of dates of symptom onset and dates of first positive specimen among participants experiencing each symptom. The fill of the bar indicates whether people with incident cases of SARS-CoV-2 first reported the symptom prior to testing positive (dark gray), the same day they tested positive (medium gray), or after testing positive (light gray)

during daily symptom diaries.  The bars depict the proportion of people with incident cases of SARS-CoV-2 who ever experienced that symptom who fell into one of these three categories. The black numbers to the right of each bar indicate the denominator for these proportions (that is, the number of people with incident cases of SARS-CoV-2 who ever experienced each particular symptom).

Table 1. Experienced symptoms and adjusted associations with daily Ct values.

| Symptom | Ever reported/asked | Symptomatic at end of follow-up* | Association with Ct value** | P value |
|---|---|---|---|---|
| Any | 62/67 | 35 | -1.6 [-2.3, -1.0] | p < 0.001 |
| **Constitutional** | | | | |
| Fatigue | 52/67 | 19 | -1.1 [-1.6, -0.6] | p < 0.001 |
| Fever | 33/67 | 5 | -1.5 [-2.3, -0.8] | p < 0.001 |
| Aches† | 21/28 | 2 | -1.3 [-2.1, -0.5] | p = 0.001 |
| Chills† | 13/28 | 1 | -2.2 [-3.4, -1.0] | p < 0.001 |
| **Gastrointestinal** | | | | |
| Diarrhea† | 10/28 | 0 | -3.0 [-4.3, -1.7] | p < 0.001 |
| Abdominal pain† | 9/28 | 0 | -0.1 [-1.5, 1.3] | p = 0.846 |
| Vomiting† | 2/28 | 0 | -0.0 [-0.1, 0.1] | p = 0.928 |
| **Lower respiratory** | | | | |
| Cough | 46/67 | 20 | -1.0 [-1.5, -0.5] | p < 0.001 |
| Chest tightness/pain | 16/67 | 2 | -2.3 [-3.4, -1.3] | p < 0.001 |
| Shortness of breath | 16/67 | 2 | -1.6 [-2.8, -0.5] | p = 0.004 |
| Wheezing | 7/67 | 1 | -2.2 [-3.5, -1.0] | p = 0.001 |
| **Upper respiratory** | | | | |
| Nasal congestion | 49/67 | 23 | -1.2 [-1.7, -0.7] | p < 0.001 |
| Runny nose | 45/67 | 15 | -1.0 [-1.6, -0.5] | p < 0.001 |
| Sore throat | 40/67 | 7 | -0.8 [-1.4, -0.2] | p = 0.008 |
| **Neurologic** | | | | |
| Headache† | 48/61 | 13 | -1.5 [-2.0, -1.0] | p < 0.001 |
| Loss of taste or smell | 29/67 | 15 | 0.2 [-0.5, 0.9] | p = 0.557 |

* Indicates the number of people who were symptomatic at the end of follow-up. The median duration of follow-up after the first positive specimen was 10 days (range: 2, 12).

** The association between symptom status and Ct values was determined by one generalized additive model per symptom, accounting for time since first testing positive, the SARS-CoV-2 test target, and the age of the case. Associations are in units of Ct values, so that negative estimates suggest that if a case reported experiencing that symptom on a particular day, the Ct values from the specimens collected that day would be lower.

†Some symptoms were assessed only at one site (Marshfield Clinic Research Institute but not Vanderbilt University Medical Center), or not assessed in children under the age of 5; see[15]

Table 2. Test characteristics among 67 incident cases

| | Overall | Age 0-11 | Age 12-17 | Age 18-49 | Age 50+ |
|---|---|---|---|---|---|
| **CDC: N1 target** | | | | | |
| Participants (n) | 30 | 6 | 5 | 14 | 5 |
| Specimens (n) | 214 | 37 | 26 | 117 | 34 |
| Ct value, median [IQR] | 28.1 [24.6, 32.9] | 30.0 [26.9, 34.5] | 28.3 [26.3, 32.4] | 27.9 [24.1, 33.3] | 27.3 [22.6, 30.3] |
| **CDC: N2 target** | | | | | |
| Participants (n) | 30 | 6 | 5 | 14 | 5 |
| Specimens (n) | 214 | 37 | 26 | 117 | 34 |
| Ct value, median [IQR] | 28.6 [25.1, 33.8] | 31.4 [27.6, 34.8] | 28.5 [26.0, 32.5] | 28.3 [24.5, 34.3] | 26.9 [22.6, 31.2] |
| **ThermoFisher: N target** | | | | | |
| Participants (n) | 37 | 12 | 6 | 13 | 6 |
| Specimens (n) | 329 | 86 | 59 | 117 | 67 |
| Ct value, median [IQR] | 22.9 [18.0, 26.9] | 24.8 [21.7, 28.6] | 25.3 [19.8, 29.3] | 21.4 [16.5, 26.7] | 19.4 [15.9, 23.9] |
| **ThermoFisher: ORF1ab target** | | | | | |
| Participants (n) | 37 | 12 | 6 | 13 | 6 |
| Specimens (n) | 327 | 83 | 59 | 118 | 67 |
| Ct value, median [IQR] | 22.4 [17.3, 26.6] | 23.9 [21.2, 27.6] | 24.8 [18.8, 28.8] | 20.6 [16.2, 26.3] | 18.8 [15.3, 23.0] |
| **ThermoFisher: S target** | | | | | |
| Participants (n) | 34 | 10 | 6 | 12 | 6 |
| Specimens (n) | 300 | 72 | 56 | 105 | 67 |
| Ct value, median [IQR] | 21.6 [17.0, 25.7] | 23.3 [20.2, 27.4] | 24.3 [18.8, 27.9] | 19.6 [15.5, 24.2] | 18.7 [15.5, 22.9] |

Table 1 shows the number of participants and number of analyzed tests for each assay target, along with the median and IQR of Ct values from those tests.

Table 3. Additional demographic variables and association with cycle threshold values.

| Predictor | Predictor level | Incident cases | Model-based association (base model)* | Model-based association (base model plus age)* |
|---|---|---|---|---|
| Vaccination | None | 60 | Reference | Reference |
| | Partial† | 7 | -1.8 [CI -2.6, -1.0] p < 0.0001 | 0.3 [CI -0.5, 1.1] p = 0.431 |
| Conditions | None | 54 | Reference | Reference |
| | At least one | 13 | -1.5 [CI -2.2, -0.9] p < 0.0001 | -0.7 [CI -1.3, -0.1] p = 0.019 |
| Sex | Female | 32 | Reference | Reference |
| | Male | 35 | -0.9 [CI -1.5, -0.4] p = 0.001 | -0.5 [CI -1.0, 0.0] p = 0.071 |

*The base model was specified similar to model 1 in the primary methods: a generalized additive model predicting cycle threshold values with a random effect spline for repeated measurements within individuals, a smoothing spline for the number of days that elapsed since the first positive specimen, and controlling for the target of each assay. The model that adjusted for age included another predictor, categorical age (0-11, 12-17, 18-49, or 50+).

†Partial vaccination was defined as having received one dose of an mRNA vaccine prior to household exposure to SARS-CoV-2. No incident cases were fully vaccinated.
